# Feasibility of Managing Diabetes Patients Identified Through Community Screening in Rural Ethiopia

**DOI:** 10.64898/2026.08.12.26360279

**Authors:** Desalegn Tsegaw Hibstu, Melaku Haile Likka, Hiwot Abera Areru, Betelhem Eshetu Birhanu, Bernt Lindtjørn

**Author notes:** Corresponding author: Desalegn Tsegaw Hibstu.

## Abstract

**Objective:** To assess the feasibility of diagnosing, enrolling, and managing adults with type 2 diabetes at the primary healthcare level using the WHO Package of Essential Non-communicable Disease Interventions (WHO-PEN) approach, following identification through community – based screening in rural Ethiopia.

**Design:** Single-arm, pre-post clinical feasibility study.

**Setting:** Primary healthcare facility in Shebedino district, Sidama National Regional State, rural Ethiopia, between September 2024 and February 2025.

**Participants:** 35 adults with newly diagnosed type 2 diabetes, identified through community-based screening and diagnosed according to the WHO and American Diabetes Association criteria.

**Interventions:** A six-month WHO-PEN based intervention comprising metformin based pharmacologic treatment, lifestyle counselling, and health system strengthening.

**Outcome measures:** Feasibility across four domains (acceptability, implementation fidelity, practicality, and preliminary clinical signal); HbA1c change from baseline to six months, analysed using the Wilcoxon signed-rank test and McNemar’s test.

**Results:** All 35 patients completed the six-month follow-up. At baseline, 6(17 %) had comorbid hypertension, 5 (14%) had elevated total cholesterol, 14 (40%) had low HDL cholesterol, 24 (69%) were underweight. Median HbA1c decreased from 52.0 mmol/mol (IQR: 50-56.0) at baseline to 43.0 mmol/mol (IQR: 42.0-46.0) at six months (reduction of 9.0 mmol/mol; p <0.001). By six months, 30 of 35 (85.7%) had HbA1c below the control threshold, versus 20 (57%) at baseline (p=0.002).

**Conclusion:** Community-identified adults with type 2 diabetes could be successfully diagnosed, linked to care, and retained using the WHO-PEN approach at a rural primary healthcare facility, with substantial improvement in glycaemic control. The absence of a control group precludes causal attribution. Medication was not adjusted for patients with persistent hyperglycaemia despite scheduled reviews, indicating a priority need to strengthen implementation of established practices rather than studying further efficacy.

**STRENGTHS AND LIMITATIONS:**

❖ This is the first clinical feasibility study to evaluate implementation of the WHO-PEN approach for diabetes management at the primary healthcare level in rural Ethiopia, assessed using a structured, multi-domain feasibility framework.
❖ Patients were identified and linked to care through a pre-established community-based screening rather than passive health facility attendance, reducing selection bias and enabling assessment among newly diagnosed adults within the community.
❖ This is among the first studies to report diabetes management outcomes in a population with a prevalence of underweight (69%), contributing evidence on diabetes care among undernourished populations in Ethiopia.
❖ The single-arm, pre-post design, conducted at a single sit without a concurrent comparison group, limits the ability to attribute
❖ The study identified implementation gaps, including the absence of medication dose adjustment for patients above the HbA1c target during follow-up, despite scheduled review visits at three and six months.

## INTRODUCTION

Diabetes mellitus is a chronic, progressive metabolic disorder and one of the most significant public health challenges of the 21st century. Worldwide, an estimated 589 million adults (11.1% of adults aged 20-79) were living with diabetes in 2024. By 2050, this number is projected to reach 852.5 million, representing a 45% increase compared with 25% global population growth over the same period [1]. Sub-Saharan Africa currently has the lowest regional diabetes prevalence (5%). However, it is expected to experience the highest percentage increase globally, a 142 % rise to 60 million by 2050, with 73% of current cases undiagnosed [1].

Diabetes causes significant morbidity, premature mortality, and economic burden. In 2019, diabetes was responsible for more than one million deaths worldwide, nearly half of which occurred among individuals younger than 70 years [2, 3]. It substantially increases the risk of lower limb amputation, neuropathy, retinopathy, stroke, cardiovascular disease, and renal failure [4–6]. In 2024, total global diabetes-related health expenditure exceeded one trillion US dollars for the first time [1].

Ethiopia faces a triple burden of communicable diseases, non-communicable diseases (NCDs), and injuries [7], with NCDS accounting for approximately 37.5% of the national disease burden [8]. The national prevalence of type 2 diabetes mellitus is estimated at 6.5%, ranging from 2% in Tigray to 14% in Dire Dawa [9]. The prevalence of undiagnosed diabetes ranges from 5.8% to 10.2% [10, 11]. A community-based study in our study area, Sidama National Regional State, reported a prevalence of 12% [12].

Diabetes risk is increased by common factors such as obesity, physical inactivity, ageing, family history, poor diet, smoking, and alcohol consumption [13, 14]. In high-income countries, the typical phenotype of type 2 diabetes is closely associated with excessive adiposity. However, a distinct and less recognised clinical phenotype sometimes referred to as lean diabetes has been described in low and middle-income countries, particularly in Sub-Saharan Africa and South Asia, where individuals of normal or low body weight can develop type 2 diabetes. The mechanisms underlying this phenotype remain incompletely understood but may involve chronic malnutrition, pancreatic beta-cell dysfunction, and altered insulin secretion [15–18].

Effective diabetes management is essential for preventing complications. Glycated haemoglobin (HbA1c), which reflects the average blood glucose levels over the preceding two to three months, is reliable indicator of long-term glycaemic control [19]; however, it remains underutilized in Ethiopia, with studies reporting limited incorporation into routine diabetes care [20–22]. Several structured care models have improved gycaemic control in type 2 diabetes. Patient-centred care models emphasis individualised treatment planning and collaborative decision-making. Evidence from randomised controlled trials and meta-analyses suggests that patient-centred self-management interventions can improve HbA1c and adherence related behaviours [23, 24].

Similarly, the Chronic Care Model (CCM) promotes proactive, team-based care through health system redesign, self-management support, clinical information systems, and regular follow-up. Interventions incorporating multiple CCM components have been associated with clinically meaningful reduction in HbA1c, particularly among individuals with poorly controlled diabetes [25]. Diabetes self-management education and support (DSMES) programmes have been shown to improve glycaemic control, medication adherence, quality of life across diverse settings [26]. Family-centred interventions and telemedicine-based strategies, including phone reminders and digital health platforms, have also demonstrated beneficial effects on HbA1c reduction [27–30]. However, implementation of these approaches often requires infrastructure, specialised personnel, and financial resources that may not be readily available in rural, resource-constrained settings.

To address NCD care in resource-limited settings, the World Health Organization developed the Package of Essential Non-Communicable Disease Interventions (WHO-PEN), a prioritised, evidence-based package of clinical protocols, essential medicines, and health system strengthening strategies designed for early detection, standardised treatment, and long-term NCD management at the primary healthcare level [31]. Ethiopia has adopted the WHO-PEN as part of its national strategy [31–33], yet its implementation in rural areas remains limited [34–36]. Building upon the WHO-PEN, the present intervention incorporated several context-specific system-strengthening components, including the establishment of a context-specific system strengthening components: dedicated NCD clinic, structured patient registry, provider training, and monthly phone-based follow-up. The detailed methodological rationale for these components has been detailed elsewhere [37].

Despite growing policy interest in decentralising diabetes care, evidence on its feasibility rural Ethiopian primary healthcare facilities remains limited. Therefore, this clinical feasibility study, linked to community-based diabetes screening, aimed to assess whether adults with newly diagnosed diabetes could be successfully diagnosed, enrolled, followed up, and managed at the primary healthcare level using the WHO-PEN approach in rural Sidama, Ethiopia.

## METHOD

### Study Design and setting

This single-arm pre-post clinical feasibility study, linked to a community-based diabetes screening, was conducted in Shebedino district, Sidama National Regional State, Ethiopia, from1September 2024 to 28 February 2025. Patients newly diagnosed with diabetes during a previously conducted community-based prevalence study were linked to care and followed at Dobe Toga Primary Health Care Unit [38].

Shebedino district is a predominantly rural, and agriculture is the principal livelihood. Ethiopia’s health system organized into three-tiers. Primary health care units, comprising health posts, health centres, and primary hospitals, provide first-line services at community and district levels. Health centres are the first point of clinical care and are staffed by health officers, nurses, midwives, and laboratory and pharmacy professionals. Health officers complete four years of degree-level clinical training and provide most outpatient diagnostic and treatment services, functioning similarly to general practitioners in higher-income settings. Patients requiring specialised care are referred to primary hospitals or higher-level facilities.

Dobe Toga Primary Health Care Unit serves the district’s catchment area, providing outpatient, laboratory, pharmacy, maternal and child health, and emergency services. Before this study, the facility did not provide organised NCD services, and suspected diabetes were referred to higher-level facilities, where distance, cost, and poor continuity of follow-up limited access to care.

### Health system strengthening and provider training

As part of the feasibility study, a dedicated NCD clinic and structured NCD patient registry were established at Dobe Toga Primary Health Care Unit.

Eight healthcare providers (four nurses, two health officers, and two laboratory professionals) completed three-day training before the intervention, delivered by a physician experienced in WHO-PEN implementation. The training covered NCD epidemiology, WHO risk assessment charts, standardised measurement techniques, diabetes management algorithms, and behavioural counselling using the 5A’s (Ask, Advise, Assess, Assist, and Arrange follow-up) and 5R’s (Relevance, Rewards, Roadblocks, and Repetition) frameworks. Practical sessions covered anthropometric measurements, glucometer use, blood sample collection, blood pressure assessment, and WHO-PEN-based case management [31].

The training physician did not return to the health centre during the six-month follow-up. Metformin doses initiated at baseline were not reviewed or titrated for any patient, including the five patients who remained above the HbA1c target throughout follow-up.

### Feasibility framework

Feasibility was assessed across four domains based on the frameworks proposed by Bowen et al [39] and Thabane et al [40]. Acceptability was assessed by enrolment rate (proportion of eligible patients who consented) and retention rate (proportion who completed all scheduled assessments at three and six months). Implementation fidelity was assessed by whether the WHO-PEN components: clinical assessment, anthropometry, blood pressure monitoring, medication review, lifestyle counselling, and monthly telephone follow-up were delivered as scheduled. Practicality was assessed by the ability to establish and sustain a dedicated NCD clinic and patient registry using existing health centre staff without specialist recruitment. A preliminary clinical outcome was assessed by change in HbA1c from baseline to six months.

### Participants

Adults aged 45 years and older with newly diagnosed type 2 diabetes were eligible for enrolment. Diabetes was diagnosed according to American Diabetes Association and World Health Organisation criteria (HbA1c ≥ 48 mmol/mol) [41,42]. Participants were eligible if they resided in the catchment area, and had no previous diabetes diagnosis or medication use, and were not pregnant.

Among the 35 patients identified through community screening, none had organ disease, cognitive impairment, or other exclusion criteria. All eligible patients were enrolled. No formal sample size calculation was performed because the primary objective was to assess the feasibility of delivering diabetes care at the primary healthcare level rather than intervention effectiveness.

### Intervention

#### Participant identification and enrolment

A community-based census was conducted in the Dobe Toga Primary Health Care catchment area among adults aged 45 years and older. Eligible participants underwent fasting blood sugar testing. Those with values >7 mmol/L received confirmatory HbA1c testing. Part pants with HbA1c ≥48 mmol/mol were diagnosed with diabetes mellitus and linked to the Dobe Toga Primary Health Care Unit for the WHO-PEN intervention. Those with elevated fasting blood sugar but HbA1c <48 mmol/mol received lifestyle counselling and were advised to continue periodic screening (Figure 1).

**Figure 1.**
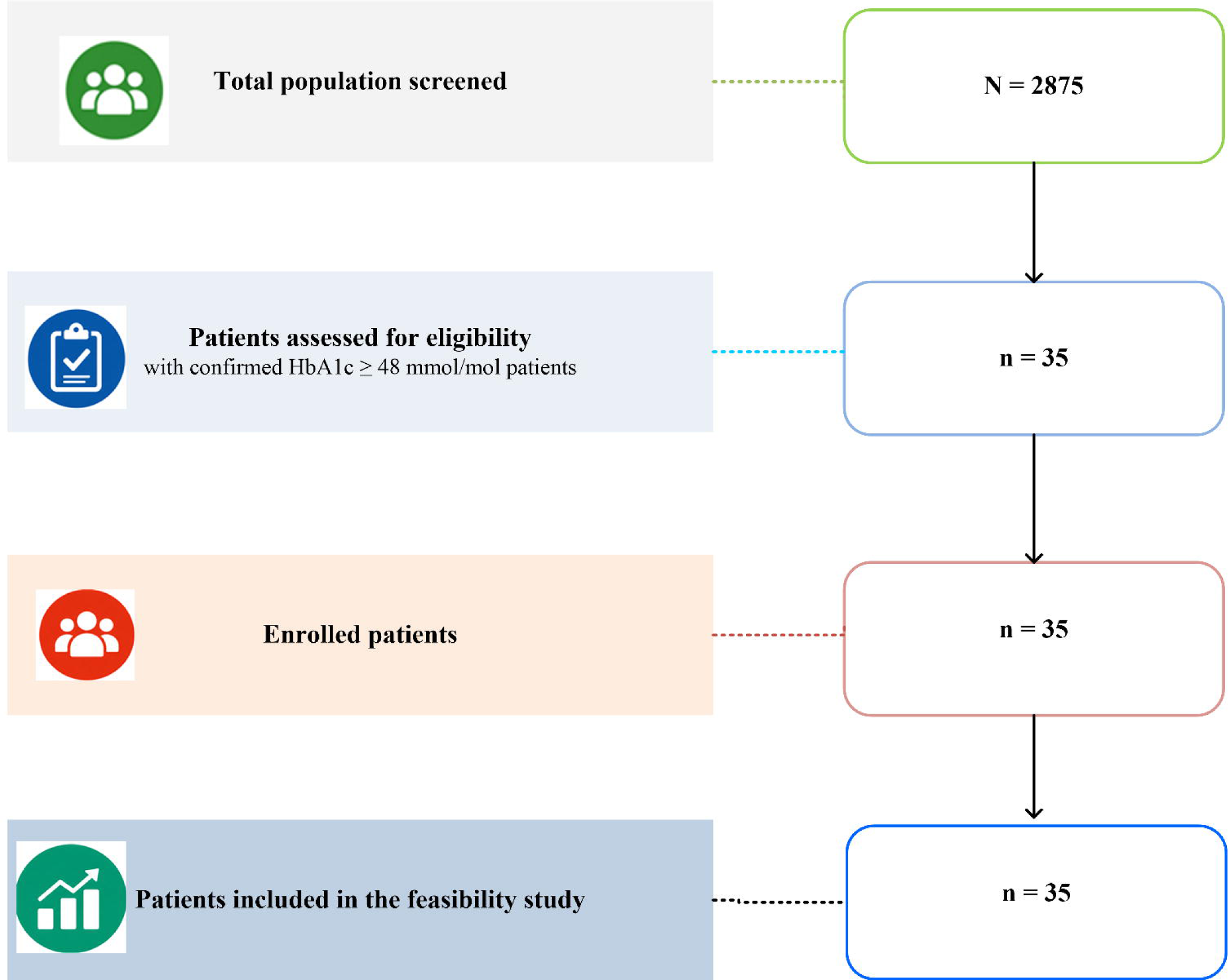

#### Clinic visits

Participants attended clinic visits at baseline, Month 3, and Month 6. Each visit included clinical assessment, anthropometric measurements, blood pressure monitoring, and lifestyle counselling according to WHO-PEN Protocol 2 [31]. Diabetes health education materials, including a banner and audio materials in the local language (*Sidaamu Affoo*) were also used.

#### Pharmacological management

Metformin was initiated for all patients at a standard dose of 500 mg daily in accordance with the WHO-PEN. The recommends medication review and adjustments at follow-up based glycaemic control; however, these adjustments were not implemented at the three or six-month visits.

#### Monthly follow-up calls

Patients received monthly structured telephone calls (10–15 minutes) from trained nurses at Dobe Toga Primary Health Care Unit. Calls followed a standardised checklist covering medication adherence. Including missed doses and side effects, dietary practices, physical activity, reminders of scheduled clinic visit, and new symptoms.

### Study Variables

The primary outcome was change in HbA1c from baseline to six months. At six months, diabetes control was defined as HbA1c < 53mmol/mol [41]. This definition was applied only to the post-treatment assessment because patients were untreated at diagnosis..

Other measurements included fasting blood sugar, systolic and diastolic blood pressure, lipid profile (total cholesterol, low-density lipoprotein cholesterol, high-density lipoprotein cholesterol, and triglycerides), anthropometric indices (body mass index and waist-to-hip ratio), and haematological parameters including red blood cell count, haemoglobin, haematocrit ,and white blood cells blood count. In this study population, anaemia was defined as haemoglobin < 12 g/dL for both males and females [43]. Socio-demographic, economic, and behavioural variables including age, sex, marital status, educational level, occupation, and distance or travel time to the health facility, wealth index, estimated annual income, tobacco use, alcohol consumption, dietary practices, and physical activity were collected at baseline to describe participant characteristics.

### Data Collection and Laboratory analysis

Data were collected at baseline, 3 months, and 6 months using a structured questionnaire adapted from the WHO-PEN clinical form [31] and WHO STEPS survey [44], translated into *Sidaamu Affoo* . The questionnaire captured socio-demographic characteristics, clinical information, medication prescriptions, and behavioural risk factors. Tobacco use, alcohol consumption, dietary practices, and physical activity were assessed at enrolment. At each visit, blood pressure, anthropometric measures (weight, waist, and hip circumference), and capillary blood glucose were measured using standardised procedures and calibrated equipment.

After an overnight fast of 8-12 hours, venous blood was collected at all three time points. Five millilitres were collected into EDTA K3 tubes for complete blood count (CBC), and 3-5 mL into serum separator tubes (SST) for lipid analysis. EDTA samples were maintained at room temperature (20–25°C) if analysed within 8 hours or refrigerated at 2–8°C for up to 48 hours. Serum was separated by centrifugation at 3000 rpm for 10 minutes and stored at 2-8 °C for up to 24 hours or at −20°C for up to 7 days when analysis was delayed.

Samples were transported daily from the Dobe Toga Primary Health Care Unit to Hawassa University Comprehensive Specialised Hospital in cold boxes maintained at 2–8°C, with transport time under 2 hours. On arrival, specimens were checked for labelling accuracy, haemolysis, clotting, lipaemia, and adequate volume; compromised samples were rejected.

Complete blood count parameters were measured using a Mindray BC-7500 haematology analyser [45]. Serum lipid parameters were analysed using a Cobas 6000 c501 chemistry analyser with automated enzymatic colourimetric assays (Roche Diagnostics, Mannheim, Germany). All laboratory analyses followed Clinical and Laboratory Standards Institute (CLSI) and International Federation of Clinical Chemistry (IFCC) recommendations [46, 47].

To minimise performance bias, healthcare providers delivering lifestyle counselling were different from those administering questionnaires and measuring outcomes. The principal investigator conducted weekly supervisory visits to monitor protocol adherence, data quality, and intervention fidelity.

### Statistical analysis

Data were analysed using Stata 17 (StataCorp, College Station, USA). Data were checked for completeness, consistency, range errors, and outliers. Normality was assessed using the Shapiro-Wilk test, histograms, and Q-Q plots. Baseline characteristics were summarised as median (IQR) for continuous variables and frequency (%) for categorical variables. The data supporting the statistical analysis is provided in Supplementary files S1.

The Wilcoxon signed-rank test was used for continuous before-and-after comparisons, and McNemar’s test for paired proportions. The chi-square was used for categorical comparisons where appropriate. The primary analysis compared baseline and six-month measurements. Given the small sample size, no subgroup or interaction analyses were pre-specified; the observations regarding the lean-diabetes phenotype and the five patients with persistent hyperglycaemia at six months are reported descriptively and should not be interpreted as formal subgroup analysis. No sensitivity analysis was undertaken because of the small sample size.

### Quality Control

Before the intervention, healthcare providers received three days of training from experienced trainer covering study procedures, data collection tools, anthropometric and blood pressure measurements, glucometer use, and structured lifestyle counselling.

Standardised equipment for anthropometry, blood pressure measurement, and blood glucose testing equipment was supplied to Dobe Toga Primary Health Care Unit. All instruments were calibrated and standardised before use.

## RESULTS

### Diagnosis and enrolment

Thirty-five patients were diagnosed with type 2 diabetes according to WHO and ADA criteria (HbA1c ≥48 mmol/mol) following the community-based screening and were enrolled in care at Dobe Toga Primary Health Care Unit.

At diagnosis, 6 patients (17%) had hypertension, 5(14%) had elevated total cholesterol, and 14 (40%) had elevated triglycerides. Twenty-four patients (69%) were underweight (BMI <18.5kg/m^2^). Clinical, nutritional, and metabolic characteristics at diagnosis are presented in Table 1.

**Table 1.** Clinical characteristics, nutritional status, and metabolic profile at diagnosis (n = 35)

| Condition | Category | n | % |
| --- | --- | --- | --- |
| Hypertension | Hypertensive | 6 | 17 |
|  | Not hypertensive | 29 | 83 |
| HbA1c at diagnosis | 48-52 mmol/mol) | 20 | 57 |
|  | ≥ 53 mmol/mol | 15 | 43 |
| HDL cholesterol | Low (≤ 1.0 mmol/L) | 14 | 40 |
|  | Normal (> 1.0 mmol/L) | 21 | 60 |
| LDL cholesterol | Elevated (≥ 2.6 mmol/L) | 8 | 23 |
|  | Normal (< 2.6 mmol/L) | 27 | 77 |
| Total cholesterol | Elevated (≥ 5.2 mmol/L) | 5 | 14 |
|  | Normal (< 5.2 mmol/L) | 30 | 86 |
| Triglycerides | Elevated (≥ 1.7 mmol/L) | 14 | 40 |
|  | Normal (< 1.7 mmol/L) | 21 | 60 |
| Nutritional status (BMI) | Underweight (< 18.5 kg/m<sup>2</sup>) | 24 | 69 |
|  | Healthy weight (18.5–24.9 kg/m<sup>2</sup>) | 10 | 29 |
|  | Overweight (≥ 25 kg/m<sup>2</sup>) | 1 | 3 |

*BMI, body mass index; HbA1c, glycated haemoglobin A1c; HDL, high density lipoprotein; LDL,* *low-density lipoprotein*

Of the 35 patients, 20 (57%) had a baseline HbA1c values of 48-52 mmol/mol, while 15 (43%) had HbA1c ≥ 53 mmol/mol. The 20 patients in the 48-52 mmol/mol range were below the post-treatment control threshold despite being newly diagnosed and untreated. This reflects the proximity of the two thresholds rather than evidence of adequate treatment or that treatment was unnecessary.

### Baseline patient characteristics

The 35 patients were predominantly female (60%, 21/35), aged 45-54 years (51%, 18/35), currently married (94%, 33/35), and without formal education (94%, 33 of 35). Detailed socio-demographic characteristics are presented in Table 2.

**Table 2.** Baseline socio-demographic and economic characteristics of patients (n = 35)

| Variable | Category | n | % |
| --- | --- | --- | --- |
| Sex | Female | 21 | 60 |
|  | Male | 14 | 40 |
| Age group (years) | 45–54 | 18 | 51 |
|  | 55–64 | 7 | 20 |
|  | 65+ | 10 | 29 |
| Marital status | Currently married | 33 | 94 |
|  | Currently not married | 2 | 6 |
| Occupation | Housewife | 18 | 51 |
|  | Farmer | 17 | 49 |
| Education | No formal education | 33 | 94 |
|  | Primary or above | 2 | 6 |
| Literacy | Cannot read or write | 31 | 89 |
|  | Can read and/or write | 4 | 11 |
| Time to health facility | ≥ 30 minutes | 31 | 89 |
|  | < 30 minutes | 4 | 11 |
| Family size | ≤ 5 members | 32 | 91 |
|  | > 5 members | 3 | 9 |
| Wealth index | Poorest | 8 | 23 |
|  | Poorer | 5 | 14 |
|  | Middle | 5 | 14 |
|  | Richer | 8 | 23 |
|  | Richest | 9 | 26 |
| Estimated annual crop production | ≤ 1 quintal | 10 | 29 |
|  | > 1 to ≤ 2 quintals | 14 | 40 |
|  | > 2 to ≤ 3 quintals | 7 | 20 |
|  | > 3 quintals | 4 | 11 |

Baseline clinical and metabolic characteristics are summarised in table 3. The Median HbA1c at diagnosis was 52.0 (50.0–56.0), median fasting blood sugar was 8.0 (7.0–9.0), median systolic and diastolic blood pressures were 135.5 and 85.0 mmHg respectively.

**Table 3.** Baseline clinical and metabolic measurements of patients (n = 35)

| Domain | Variable | Median (IQR) |
| --- | --- | --- |
| Glucose level | HbA1c (mmol/mol) | 52.0 (50.0–56.0) |
|  | Fasting blood sugar (mmol/L) | 8.0 (7.0–9.0) |
| Blood pressure | Systolic BP (mmHg) | 135.0 (134.0–137.0) |
|  | Diastolic BP (mmHg) | 85.0 (84.0–87.0) |
| Lipid profile | HDL cholesterol (mmol/L) | 0.91 (0.83–1.06) |
|  | LDL cholesterol (mmol/L) | 2.19 (1.94–2.60) |
|  | Total cholesterol (mmol/L) | 3.99 (3.88–4.65) |
|  | Triglycerides (mmol/L) | 1.69 (1.64–2.03) |
| Haematologic test | Haemoglobin (g/dL) | 11.0 (11.0–12.0) |
| Anthropometric | BMI (kg/m<sup>2</sup>) | 17.7 (16.6–18.7) |
|  | Weight (kg) | 44.0 (40.0–48.0) |
|  | Waist circumference (cm) | 68.0 (67.5–72.0) |

*BMI, body mass index; BP, blood pressure; HbA1c, glycated haemoglobin A1c; HDL, high* *density lipoprotein; IQR, interquartile range; LDL, low-density lipoprotein*

### Implementation Fidelity: Treatment delivery

All 35 patients received metformin as first-line pharmacologic therapy, in accordance with the WHO-PEN protocol. Twelve patients (34%) had borderline HbA1c values of 48-50 mmol/mol at diagnosis; metformin was nevertheless initiated for all confirmed diabetes cases to assess implementation of the standard WHO-PEN treatment pathway.

All patients received structured lifestyle counselling according to WHO-PEN Protocol 2 [31]. Patients with hypertension (6,17.1%)) received additional cardiovascular risk counselling. No lipid-lowering medications were prescribed during the study period.

### Acceptability: Retention and follow-up

All the 35 patients completed scheduled follow-up visits at three and six months, resulting in 100% retention. No patients were lost to follow-up, transferred, or withdrew. All planned clinical assessments, laboratory investigations, and HbA1c measurements were completed at follow-up.

### Preliminary signal of clinical benefit: Treatment outcomes

Median HbA1c declined from 52.0 mmol/mol (IQR: 50.0-56.00) at baseline to 43.0 mmol/mol (IQR: 42.0-46.0) at six month, a median reduction of 9.0 mmol/mol (95% CI: 7.0-12.0; Z = 5.217, p < 0.001). Median fasting blood sugar declined from 8mmol/L to 7.0mmol/L.

Five patients (14.3%) had HbA1c ≥ 53 mmol/mol) at six months. Medication review was not performed at the three-month visit for any patient.

Blood pressure also improved, with median systolic and diastolic blood pressure decreasing by 10 and 7 mmHg a, respectively. Hypertension prevalence declined from 17% (6/35) to 6% (2/35) at six months, although this reduction was not statistically significant (p = 0.125). Changes in other clinical measurements are presented in Table 4.

**Table 4.**
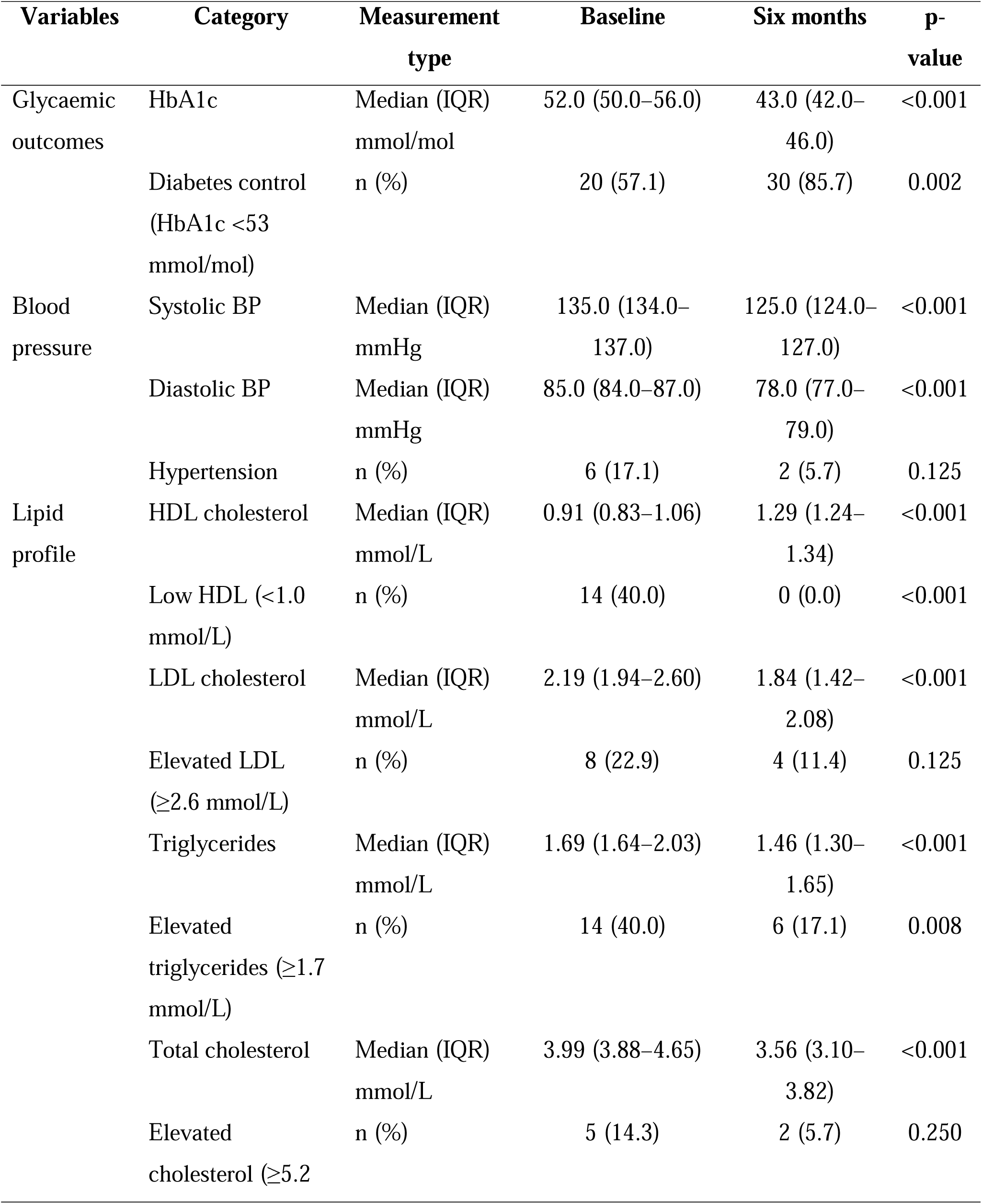
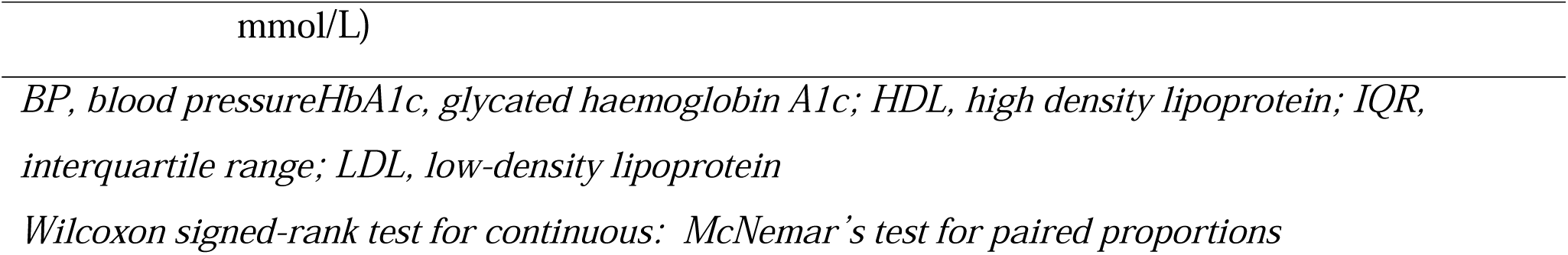
Changes in Clinical measurements: continuous values and categorical proportions, baseline versus six Months (n = 35)

### Practicality: Health centre function and implementation gaps

The same WHO-PEN trained health officers provided care throughout follow-up. The dedicated NCD clinic, patient registry, scheduled clinic visits, lifestyle counselling, and monthly follow-up calls were maintained throughout the study period.

An important implementation gap concerned medication management. Five patients (14%) remained above the HbA1c target at six months, yet no medication adjustments, including dose escalation, addition of a second agent, or therapeutic switch, were made at either the three-month or six-month visits. All patients therefore remained on the initial metformin regimen throughout the study.

## DISCUSSION

This study showed that adults with type 2 diabetes identified through community screening could be linked to care and retained in follow-up at a rural Ethiopian primary healthcare facility that previously had no NCD services. The proportion of participants with HbA1c below the post-treatment control threshold increased from 57.1 % at baseline to 85.7% at six months. However, no medication adjustments were made for patients who remained above the glycaemic target, highlighting an important gap in treatment individualisation during routine care.

The baseline HbA1c classification requires careful interpretation. Although 57.1% patients were below the post-treatment control threshold of < 53mmol/mol, all 20 had HbA1c values of 48-52 mmol/mol and were newly diagnosed and untreated. They therefore met the diagnostic threshold for diabetes (≥48 mmol/mol) while remaining below the threshold for poor glycaemic control. This classification reflects the proximity of the two thresholds rather than adequate diabetes management or an indication that treatment was unnecessary.

The observed improvement in glycaemic in this feasibility study is broadly consistent with evidence from primary care-based diabetes LMIC settings. Studies from Gaza and Palestine reported improved glycaemic measures among patients receiving WHO-PEN-based care compared with the conventional care [48, 49]. Although these studies differ in design and outcomes, they provide supporting evidence for the potential value of structured primary care approaches for diabetes management.

In Ethiopia, a pre-post study of nurse-led structured diabetes management in Tigray also demonstrated significant HbA1c improvement [50]. Unlike this intervention, which focused mainly on nurse-led education, the present model combined lifestyle counselling, pharmacological management, patient follow-up, and basic health system strengthening.

A systematic review of primary care-based diabetes interventions in LMICS reported HbA1c reductions across diverse healthcare settings, with effective approaches commonly involving case management, community health workers, and structured follow-up [51]. A quasi-experimental study in South Africa introducing group diabetes education as part of intensified care also reported glycaemic improvements in glycaemic control [52]. However, the intervention differed from the present study, by focusing on group education rather than combining individual counselling, medication management, and health system strengthening. A randomised controlled trial from Rwanda reported a larger reduction in HbA1c than observed in this study [53]. The difference may reflect longer follow-up duration, baseline HbA1c levels, intervention intensity, and study design

The median reduction of 9 mmol/mol should, however, be interpreted as a preliminary feasibility finding rather than evidence of treatment effectiveness. Regression to the mean may have contributed because participants were identified at or near the diagnostic threshold. Newly diagnosed patients may also improve following diagnosis, treatment initiation, and increased clinical attention. Increased monitoring and contact may additionally have produced a Hawthorne effect. The Pre-post design without a comparison group cannot distinguish these effects from the contribution of the intervention. The concurrent improvement in fasting blood sugar supports the observed improvement in glycaemic status but does not establish a pharmacological effect [54].

A notable characteristic of this cohort was the high prevalence of undernutrition: 69% of participants were underweight at enrollment. This finding is consistent with reports describing lean diabetes phenotypes in parts of Sub-Saharan Africa, although this study was not designed to classify diabetes subtypes [15]. Because much diabetes management evidence has been generated in populations with higher levels of overweight and obesity, its applicability to underweight populations requires careful consideration. Available evidence suggests metformin can remain effective among lean individuals [55], and the improvement observed in this cohort is consistent with this evidence. Future studies should assess whether diabetes management and nutritional counseling require adaptation for population with a high burden of undernutrition.

An important implementation finding was that five participants remained above the HbA1c target at six months, yet their treatment regimens were not modified during follow-up. This suggests limited individualisation of treatment decision rather than failure of the WHO-PEN approach itself. Health officers received a single training session without subsequent clinical mentorship, and the training physician did not return during follow-up. These findings suggest that initial training may be insufficient to support sustained treatment optimization in rural primary care settings.

The successful establishment of a dedicated NCD clinic, implementation of a patient registry, and integration of diabetes follow-up within a facility that previously provided no NCD care demonstrate that WHO-PEN can be operationalized in rural Ethiopian primary healthcare with basic health system strengthening support. Complete patient retention during six months of follow-up further suggests that the model was acceptable patients and feasible to deliver. However, these findings should be interpreted as evidence of implementation feasibility rather effectiveness.

The study has several limitations. The small sample size limits statistical precision, the single-site design limits generalisability, and the six-month follow-up limits assessment of long-term sustainability. The absence of a control group prevents causal attribution of changes in HbA1c to the intervention. The lack of ongoing clinical mentorship was both a limitation of the implementation model and an important finding for future scale-up.

These findings have implications for rural primary healthcare. The WHO-PEN-based diabetes services can be established in facilities with limited prior NCD capacity when supported by provider training, patient registries, and structured follow-up. However, service establishment alone may not ensure treatment optimisation; ongoing clinical support is needed to promote individualised treatment decisions. Given the existing evidence from multiple settings worldwide that standard diabetes management practices are effective, these findings highlight the need how established diabetes care practices can be implemented and sustained in resource limited rural settings, including whether nutritional assessment and counseling should be adapted for populations with high levels of undernutrition.

## CONCLUSION AND RECOMMENDATION

This clinical feasibility study demonstrated that adults with diabetes identified through community screening can be successfully diagnosed, linked to care, and retained in follow-up at a rural Ethiopian primary healthcare facility using the WHO-PEN approach. Glycaemic control improved over six months, but medication was not adjusted for patients who remained above the HbA1c target despite scheduled follow-up visits. This finding highlights an important implementation gap and suggests that establishing WHO-PEN services may not ensure treatment optimisation.

These findings support the feasibility of integrated diabetes care into rural primary healthcare through targeted health system strengthening, including dedicated NCD clinics, patient registries, provider training, and structured follow-up. Given the strong evidence from a wide range of settings that standard diabetes management practices are effective, the priority in this setting should be to strengthen their implementation, particularly by ensuring that scheduled review visits result in appropriate medication review and adjustment for patients who remain above target. The high prevalence of undernutrition further suggests that future implementation efforts should incorporate nutritional assessment and context-adapted counselling for patients with undernutrition.

## DECLARATIONS

## Acknowledgements

The authors gratefully acknowledge the Government of Norway for funding through the NORHED SENUPH-II programme. We thank the Shebedino district administration, data collectors, and study participants for their support. We sincerely thank the health professionals at Dobe Toga Primary Health Care Unity for delivering the interventions as intended, the laboratory staff at Hawassa University Comprehensive Specialized Hospital for their technical support in sample analysis

## Contributors

DTH, MHL, HAA, BEB and BL were involved in the study conception, data collection, statistical analysis, investigation, methodology development, project coordination, software application, supervision, data validation, and visualization of findings. All authors drafted and revised the manuscript. All authors approved the final version. DTH is the guarantor.

## Patient and Public involvement

Neither patients nor members of the public were involved in the design, execution, reporting, or dissemination of this research

## Patient consent for publication

Not applicable

## Ethics approval

This research involved human participants and received ethical clearance from the Institutional Review Board Hawassa University, College of Medicine and Health Sciences (Ref. no. IRB/100/16, issued on 12 March 2024). Additional administrative approval was obtained from the Sidama National Regional State Public Health Institute. Permission was granted by the Shebedino district health office and kebele administrators. Written informed consent was secured from every participant prior to their enrolment in the study.

## Data availability statement

All data supporting the findings of this study are openly available as supplementary material, including the anonymised data set (S1), study flow diagram (Figure 1), and the STROBE checklists (S2 and S3). All supplementary files can be accessed with this manuscript submission.

